# Gene-corrected human iPSC-derived cardiomyocytes and skeletal muscles reveal partial dystrophin Dp427 preservation and cardiac Dp116 expression in Duchenne muscular dystrophy

**DOI:** 10.1101/2025.07.11.25331067

**Authors:** Marta Białobrzeska, Marta Przymuszała, Anna Potulska-Chromik, Anna Kostera-Pruszczyk, Jacek Stępniewski, Urszula Florczyk-Soluch, Józef Dulak

**Author notes:** First co-authors contributed equally to the present work (Marta Białobrzeska, Marta Przymuszała).

## Abstract

**Background:** Duchenne muscular dystrophy (DMD) is a severe X-linked neuromuscular disease caused by mutations in the *DMD* gene, leading to the absence or dysfunction of dystrophin. While cardiac and skeletal muscles are both affected, tissue-specific differences in disease manifestation and dystrophin regulation remain poorly understood.

**Methods:** To investigate these differences, we established a human induced pluripotent stem cell (hiPSC) model of DMD from peripheral blood mononuclear cells (PBMC) of a patient carrying a splice-site mutation in intron 68 (c.9975-1G>T). An isogenic control line was generated via CRISPR/Cas9 correction. Both repaired and DMD hiPSCs were differentiated into cardiomyocytes (hiPSC-CMs) and skeletal muscle cells (hiPSC-SMs). Transcript and protein analyses were performed, along with functional assessment using microelectrode array.

**Results:** Transcript analysis revealed an in-frame deletion of two amino acids (Tyr3325 and Arg3326) due to skipping of the first six nucleotides of exon 69. Despite this, near full-length Dp427 was detected by western blot, along with expression of Dp116 in hiPSC-CMs. Dystrophin levels were preserved in DMD hiPSC-CMs but markedly reduced in hiPSC-SMs, suggesting tissue-specific regulation. Functional analysis showed altered β-adrenergic responsiveness in DMD hiPSC-CMs, with increased beating frequency and accelerated repolarization upon isoproterenol stimulation.

**Conclusions:** Our study identifies a splice-site mutation that preserves high level of dystrophin expression in cardiac but reduced in skeletal muscle and reveals Dp116 expression in cardiomyocytes. These findings highlight the importance of tissue context in DMD and demonstrate the power of hiPSC-based systems for dissecting mutation-specific effects.

## 1. Introduction

Duchenne muscular dystrophy (DMD) is an X-linked neuromuscular disorder that affects approximately 1 in 5000 boys ^1^. It is caused by mutations in the *DMD* gene on Xp21.2, which encodes dystrophin – a 427 kDa cytoskeletal protein essential for maintaining sarcolemmal integrity during muscle contraction ^2^. DMD is the most common form of muscular dystrophy, with symptoms typically appearing between the ages of 1 and 3 and including delayed motor milestones, Gowers’ sign, and pseudohypertrophy of the calves. In early adolescence, progressive muscle wasting causes loss of ambulation, and by the second to fourth decade of life, cardiomyopathy and respiratory failure are the leading causes of patients’ death ^3,4^.

The *DMD* gene is the largest known in the human genome spanning approximately 2.4 million base pairs and comprising 79 exons. The gene contains at least seven independent promoters ^5^. Alternative promoter usage and extensive splicing give rise to at least eight tissue-specific isoforms, including full-length Dp427m expressed in skeletal and cardiac muscle, and shorter isoforms such as Dp116 in Schwann cells ^6^. Dp116 transcript was also identified in adult cardiac muscle, and has been suggested to play a role in DMD-related cardiomyopathy ^7^. The type and location of the mutation strongly influences the clinical severity of DMD ^1,6^. Most cases are caused by out-of-frame mutations that disrupt open reading frame (ORF), resulting in premature stop codons, truncated transcripts, and complete dystrophin loss. In contrast, in-frame mutations, typically preserve ORF and produce truncated, but partially functional dystrophin, resulting in milder clinical phenotype observed in Becker muscular dystrophy (BMD) ^1,6^.

Dystrophin is a key component of the dystrophin-associated glycoprotein complex (DAGC), a transmembrane protein network that connects the intracellular cytoskeleton to the extracellular matrix (ECM) via laminin-211. This connection is essential for sarcolemma stability and mechanical protection of muscle fibers during contraction ^8^. The DAGC is composed of extracellular components (α-dystroglycan and laminin-211), transmembrane proteins (β-dystroglycan, α-, β-, γ-, and δ-sarcoglycans, and sarcospan), and intracellular adaptor proteins (α1- and β1-syntrophin, α-dystrobrevin, and neuronal nitric oxide synthase, nNOS) ^9^. In DMD, loss of DAGC destabilizes the sarcolemma, increasing its susceptibility to mechanical damage ^10^. Although dystrophin deficiency leads to the depletion of most DAGC components in skeletal muscle, cardiac muscle retains a majority of these proteins, which may partially compensate for the structural deficit ^10^. Nevertheless, membrane instability is evident in dystrophic hearts through elevated serum biomarkers such as creatine kinase (CK), calcium overload ^11,12^, and subsequent disturbances in iron homeostasis ^13^, mitochondrial function ^14^, and reactive oxygen species (ROS) production ^15,16^.

Access to primary muscle tissue from patients as an investigative material is highly limited especially in the case of cardiac muscle, where biopsies are rarely performed due to their invasive nature. Therefore, human induced pluripotent stem cell (hiPSC) technology, when combined with Clustered Regularly Interspaced Short Palindromic Repeats and CRISPR-associated protein 9 (CRISPR-Cas9) gene editing system, provides a powerful platform for *in vitro* genetic disease modelling, preserving patient’s genetic background ^17,18^. This approach is particularly valuable in the context of DMD, which affects both skeletal and cardiac muscles.

Herein, a novel mutation in 3’ splice site of intron 68 (c.9975-1G>T) in *DMD* was identified in a young boy through whole-exome sequencing (WES). This mutation led to the excision of the first six nucleotides of exon 69, resulting in the deletion of two amino acids, tyrosine and arginine (positions 3325 and 3326), while maintaining the reading frame and dystrophin expression.

## 2. Materials and methods

**A full description of the methods is provided in the Supplemental Material.**

### 2.1 Patient

The patient described in this study was a boy with no family history of neuromuscular disorders, who presented initial symptoms in early childhood, including mild difficulty climbing the stairs and markedly elevated CK levels (17 300 U/l; reference value: <163 U/l). In early childhood, a de novo c.9975-1G>T mutation in intron 68 of the *DMD* gene was identified by whole-exome sequencing (WES). Upon diagnosis, the boy exhibited hyper lordosis and calf hypertrophy, a North Star Ambulatory Assessment (NSAA) score of 19/34, and a 6-minute walk test (6MWT) distance of 325 meters (normal range: 400–700 meters). Overall, the clinical presentation during described developmental stage was consistent with a mild DMD phenotype.

### 2.2 hiPSCs culture

The study was approved by the Bioethical Committee of the Warsaw Medical University (approval no. KB/111/2019, approved on 10.06.2019; project title: Molecular mechanisms of heart failure in Duchenne and Becker muscular dystrophy). hiPSCs were generated from DMD patient peripheral blood mononuclear cells (PBMCs) using CytoTune™-iPS 2.0 Sendai Reprogramming Kit (ThermoFisher Scientific) according to the manufacturer’s protocol as reported by us previously ^19^.

### 2.3 CRISPR-mediated genome editing

Single-guide RNAs (sgRNAs) targeting the mutated region of the *DMD* gene were designed using the CHOPCHOP tool and cloned into pSpCas9(BB)-2A-Puro vector (Addgene #62988). For precise gene correction, a homology-directed repair (HDR) template was constructed and cloned into the pcDNA3.1/Hygro(+) plasmid (Invitrogen #V87020). The HDR template contained two homology arms flanking the mutated region and included silent PAM-disrupting mutations to prevent re-cutting by Cas9. All constructs were verified by Sanger sequencing.

### 2.4 Cardiac differentiation

hiPSCs were differentiated into CMs according to the established cardiac differentiation method ^20,21^ which involves the use of small molecules modulating Wnt/β-catenin pathway. Previously described DMB03 isogenic lines, generated from PBMCs of the patient with deletion of *DMD* exons 48-50, ^13^ have been used as the controls to demonstrate the presence and absence of dystrophin in repaired and DMD hiPSC-CMs.

### 2.5 Skeletal muscle differentiation

hiPSCs were differentiated into skeletal muscle cells (hiPSC-SMs) using the Skeletal Muscle Differentiation Kit (AMSBIO) according to the manufacturer’s protocol.

## 3. Results

### 3.1 CRISPR/Cas9-mediated generation of repaired hiPSCs

The *DMD* c.9975-1G>T mutation in patient-derived hiPSCs was repaired using a CRISPR/Cas9-mediated homology-directed repair (HDR) strategy (Fig. 1A). We designed two sgRNAs targeting *DMD* gene and cloned into pSpCas9(BB)-P2A-Puro vector. Plasmids were nucleofected into hiPSCs and selected with puromycin. Surveyor nuclease assay on isolated genomic DNA revealed genome editing efficiency. Two additional bands appeared in cells transfected with sgRNA2, indicating sequence alterations at the target site (Fig. 1B). The correction strategy involved a repair template containing left (LHA) and right (RHA) homology arms spanning introns 68 and 69 and exon 69 (Fig. 1C). Homology arms were PCR-amplified (Supplementary Material, Fig. S1) and assembled by overlap extension PCR (Fig 1C, Supplementary data, Fig. S1). Repair template assembly was confirmed by Sanger sequencing (Fig. 1C). Separate plasmids encoding Cas9/sgRNA and the repair template were co-delivered into hiPSCs, promoting HDR-mediated repair of DMD c.9975-1G>T mutation. Clonal populations were screened using the Surveyor nuclease assay, and clones exhibiting cleavage products indicative of correction were selected for Sanger sequencing. Representative results are shown in Supplementary Material, Fig. S2. Successful correction of the c.9975-1G>T mutation was confirmed in clone 10.

**Fig. 1.**
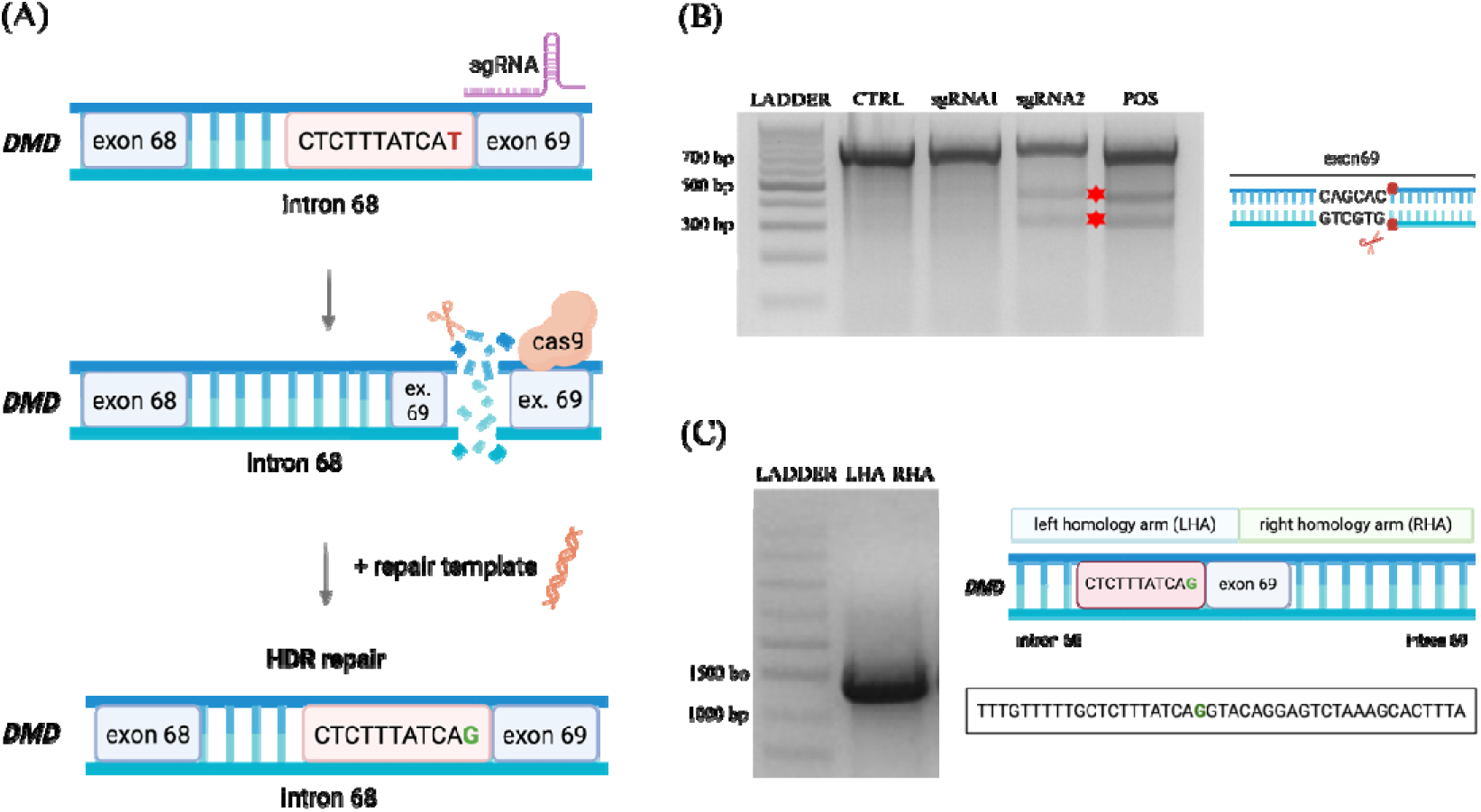
CRISPR/Cas9-mediated correction of the *DMD* mutation in hiPSCs. **(A)** Genome editing strategy that utilizes single-guide RNA (sgRNA) and Cas9 complex targeting specific site near *DMD* c.9975-1G>T mutation. **(B)** Genomic DNA from DMD human induced pluripotent stem cells (hiPSCs) transfected with pSpCas9(BB)-2A-Puro encoding sgRNA1 or sgRNA2 was analyzed by Surveyor nuclease assay. Cleavage products, indicative of indel formation, were detected only in sgRNA2 (258 bp and 501 bp), but not in sgRNA1 or the untreated control (CTRL). A positive control (POS) confirmed assay’s functionality. The schematic on the right illustrates the sgRNA target site and predicted Cas9 cleavage site within the amplified region. Ladder: DNA Marker 1 (100-1000 bp). **(C)** The repair template designed for DMD hiPSCs included two homology arms to enable gene correction via homology-directed repair (HDR). Left: analysis of overlap extension PCR product (1268 bp). Inset: sequencing result of the repair template confirming the presence of the correct nucleotide G at the site of c.9975-1G>T mutation. Ladder: IDEAL2 DNA Ladder (700–9276 bp).

### 3.2 Characteristic of hiPSCs

To assess the efficiency of CRISPR/Cas9-mediated correction of c.9975-1G>T mutation in the *DMD* gene (Fig. 2A), we first performed a Surveyor nuclease assay on genomic DNA extracted from nucleofected hiPSCs. This preliminary screening indicated successful editing events (Supplementary data, Fig.S2). Sanger sequencing verified the correction of the c.9975-1 mutation, showing the presence of a T nucleotide in the repaired clone, whereas the DMD line retained the G nucleotide (Fig. 2C). Both the repaired and DMD hiPSC lines were further subjected to G-banding analysis, confirming normal karyotypes (Fig. 2B). To evaluate whether genome editing affected pluripotency, immunofluorescence staining was performed. The repaired and DMD hiPSCs expressed key pluripotency markers, including NANOG, OCT3/4, and SSEA4 (Fig. 2D). Upon spontaneous differentiation via embryoid body formation, cells expressed markers characteristic of all three germ layers: GATA4 (endoderm/mesoderm), neurofilament heavy chain (NFH) (ectoderm), and vimentin (mesoderm) (Fig. 2E).

**Fig. 2.**
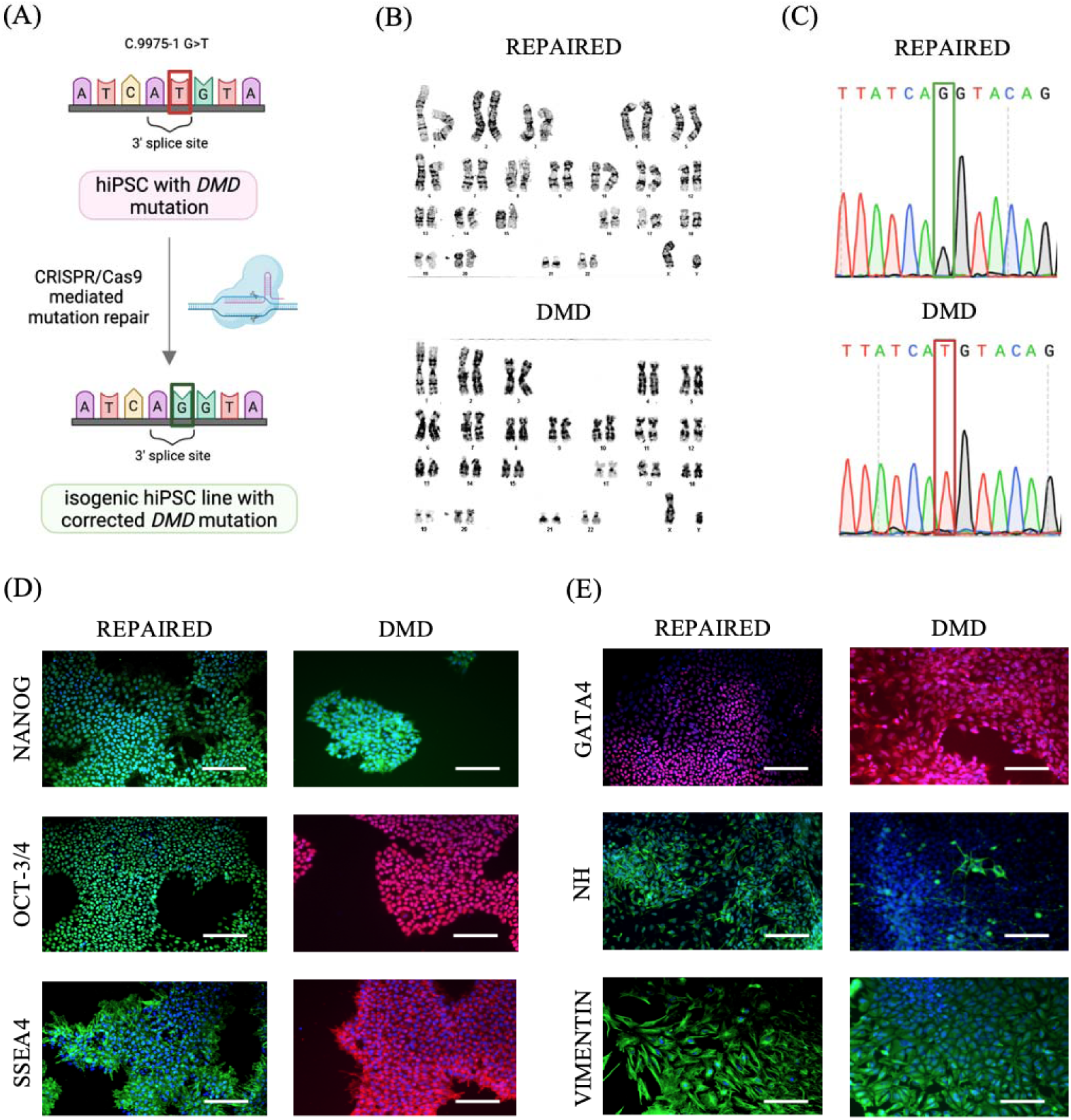
Characterization of repaired and DMD hiPSC lines. **(A)** Comparison of the DNA sequence at c.9975-1 site between gene-repaired (repaired) human induced pluripotent stem cells (hiPSCs) (G) and unrepaired DMD hiPSCs (T). **(B)** Karyotypic analysis of repaired and DMD hiPSC lines. **(C)** Representative chromatograms from Sanger sequencing of gene-repaired and DMD hiPSC lines. The point mutation present in the DMD line is repaired in the repaired line, as indicated by the restored wild-type sequence. **(D)** Immunocytochemistry staining for pluripotency markers NANOG, OCT-3/4 and SSEA4. Scale bars: 100 μm. **(E)** Immunofluorescent staining for endo-(GATA4), ecto-(neurofilament heavy chain – NH) and mesoderm (GATA4, vimentin) markers after spontaneous differentiation. Scale bars: 100 μm.

### 3.3 Cardiac and skeletal differentiations

To investigate the functional consequences of a *DMD* point mutation in disease-relevant tissues, we applied both cardiac and skeletal muscle differentiation protocols. Given that dystrophin is predominantly expressed in striated muscle, this strategy enabled the assessment of DMD-associated phenotypes, and following gene repair, of hiPSC-CMs and hiPSC-SMs derived from the same genetic background.

Cardiomyocyte differentiation was carried out using a monolayer-based protocol involving sequential modulation of WNT signalling pathway (Fig. 3A). Spontaneous contractions were first observed between days 7 and 10 in both groups. By day 25, cells established structured, beating monolayers, representative of which are shown in the bright-field images in Fig. 3B. The use of metabolic selection during differentiation led to effective enrichment of cardiomyocytes, reflected by a marked reduction in non-contractile cell types. At this stage, no clear morphological differences were observed between repaired and DMD hiPSC-CMs. Both exhibited expression of key cardiac markers, including α-actinin (ACTN2) and troponin I (TnI), as confirmed by immunofluorescence analysis (Fig. 3C). In addition, well-organized sarcomeric structures were evident (Fig. 3C, frames A and B), consistent with successful cardiomyocyte differentiation.

**Fig. 3.**
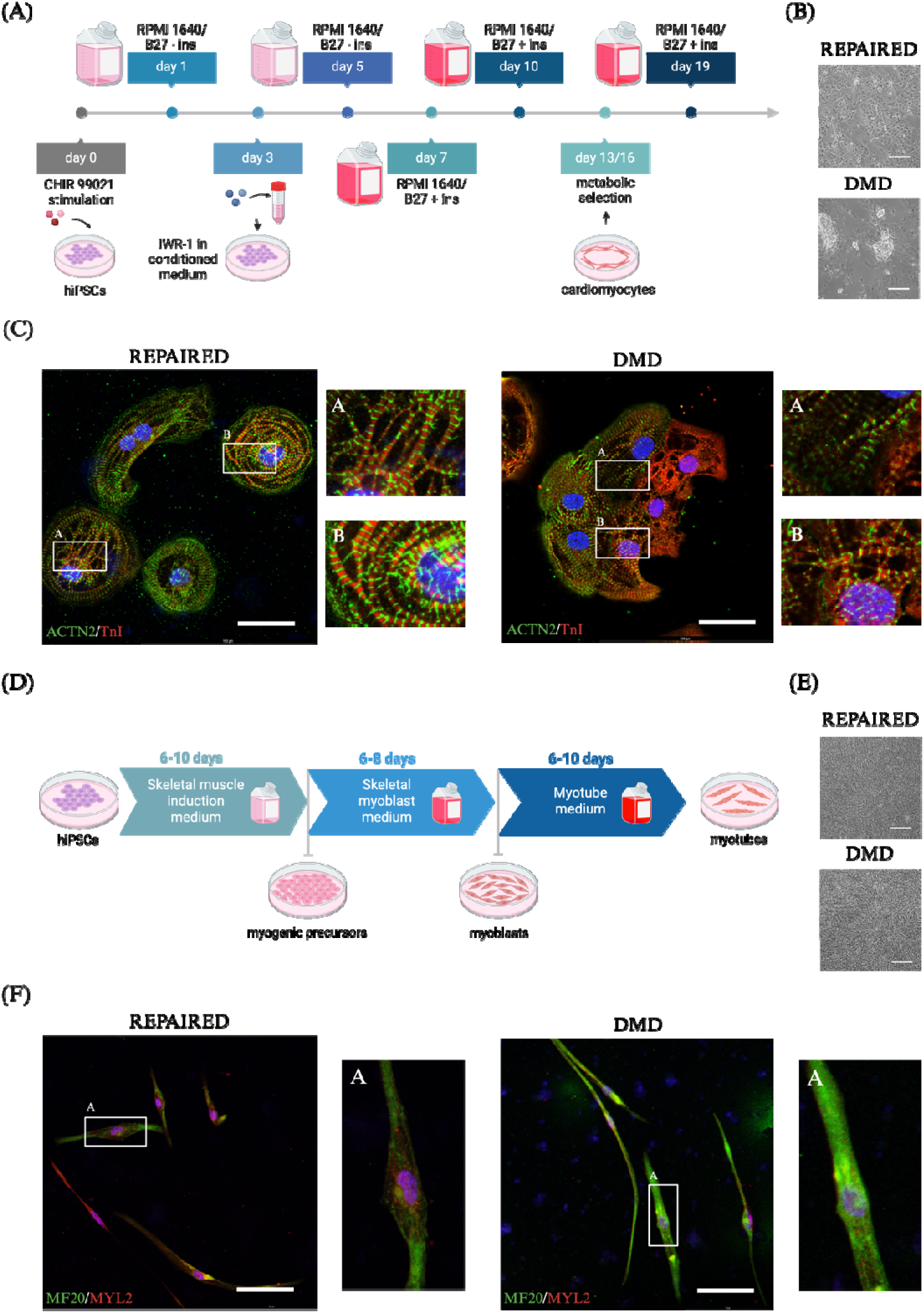
Differentiation of hiPSCs to cardiomyocytes and skeletal muscle cells. **(A)** Scheme of cardiac differentiation based on Wnt/β-catenin pathway modulation. **(B)** Representative bright-field images of repaired and DMD hiPSC-CMs on day 25 of differentiation, showing monolayer morphology. Scale bar: 200 μm. **(C)** Immunofluorescent (IF) images of hiPSC-CMs stained for cardiac markers: α-actinin (ACTN2, green), troponin I (TnI, red), nuclei stained with Hoechst (blue). Scale bars: 100 µm. **(D)** Scheme of skeletal muscle differentiation using a media-based protocol. **(E)** Representative bright-field images of repaired and DMD hiPSC-SMs. Scale bars: 200 μm. **(F)** IF images of hiPSC-SMs stained for cardiac markers: myosin heavy chain (MF20, green), myosin light chain 2 (MYL2, red), nuclei stained with Hoechst (blue). Scale bars: 100 µm. hiPSC-CMs – human induced pluripotent stem cell-derived cardiomyocytes, hiPSC-SMs - human induced pluripotent stem cell-derived skeletal muscles.

Skeletal muscle differentiation was achieved using a media-based protocol, which directed hiPSCs through sequential stages of myogenic commitment, including progenitor cells, myoblasts, and mature myotubes (Fig. 3D). Early-stage cultures were characterized by oval-shaped, rapidly proliferating cells, which, upon reseeding at defined density, gave rise to spindle-shaped myoblasts exhibiting reduced proliferative capacity. These myoblasts subsequently fused to form elongated, multinucleated myotubes, as shown in Fig. 3E. Differentiation towards the myogenic lineage was further confirmed by the expression of myosin light chain (MYL2) and myosin heavy chain (MF20), two established markers of mature muscle cells (Fig. 3F).

### 3.4 Differences in expression levels of the dystrophin in hiPSC-CMs and hiPSC-SMs

The patient’s mutation is located in the last nucleotide of intron 68, within the 3’ splice acceptor site. It was expected to cause aberrant splicing, leading to exon 69 skipping, introduction of a premature stop codon, and ultimately the absence of dystrophin expression. Unexpectedly, transcript analysis revealed the inclusion of exon 69 in dystrophin mRNA from both repaired and DMD hiPSC-CMs and hiPSC-SMs (Supplementary data, Fig. S3). However, sequencing of dystrophin mRNA demonstrated that the point mutation caused an excision of the first six nucleotides of exon 69 (Fig. 4B), resulting in the deletion of two amino acids, tyrosine and arginine (positions 3325 and 3326), while preserving the reading frame.

**Fig. 4.**
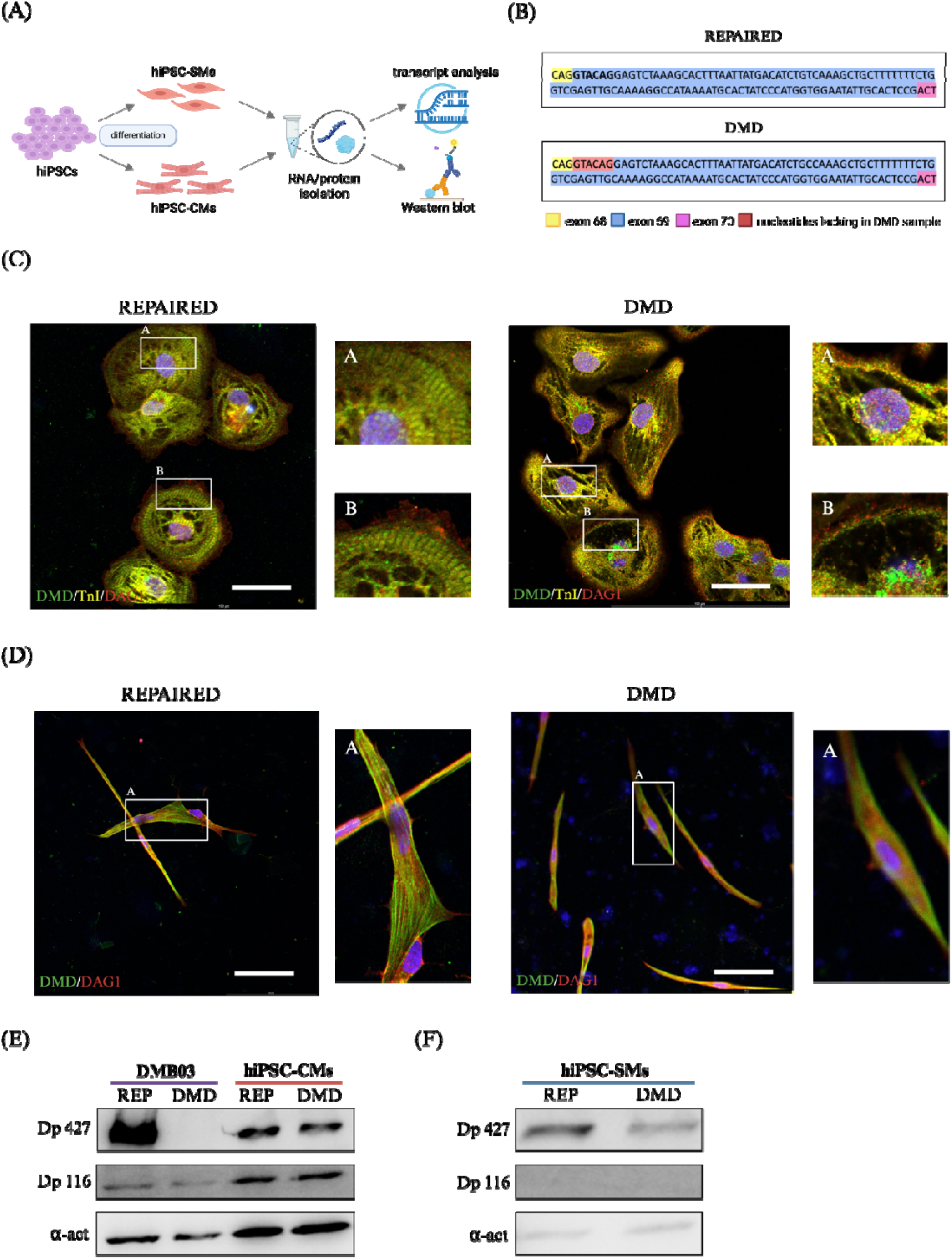
Expression of dystrophin and dystroglycan in hiPSC-CMs and hiPSC-SMs. **(A)** Schematic overview of the experimental design: hiPSCs derived from a repaired and DMD patient lines were differentiated into hiPSC-CMs and hiPSC-SMs, followed by RNA and protein analysis. **(B)** Sanger sequencing of dystrophin mRNA revealed full-length inclusion of exon 69 in the repaired line, whereas the DMD transcript lacked the first six nucleotides of exon 69. **(C)** Immunofluorescent (IF) images of hiPSC-CMs derived from repaired and DMD lines stained for dystrophin (DMD, green), troponin I (TnI, yellow), dystroglycan (DAG1, red) nuclei stained with Hoechst (blue). Scale bars: 100 µm. **(D)** IF images of hiPSC-SMs derived from repaired and DMD lines stained for dystrophin (DMD, green), dystroglycan (DAG1, red), nuclei stained with Hoechst (blue). Scale bars: 100 µm. **(E)** Representative western blot analysis of dystrophin isoforms Dp427 and Dp116 in repaired (REP) and DMD hiPSC-CMs. Dystrophin is absent in hiPSC-CMs generated from DMB03 DMD hiPSCs. α-actinin was used as loading control. **(F)** Representative western blot analysis of dystrophin isoforms Dp427 and Dp116 in repaired (REP) and DMD hiPSC-SMs. α-actinin was used as loading control. hiPSC-CMs – human induced pluripotent stem cell-derived cardiomyocytes, hiPSC-SMs - human induced pluripotent stem cell-derived skeletal muscles.

Confocal microscopy revealed discontinuities or membrane irregularities in DMD hiPSC-derived cardiomyocytes (hiPSC-CMs), which were absent in the repaired cells (Fig. 4C). These defects were visible in both DMD and dystroglycan (DAG1) immunostaining (Fig. 4C, frames A and B), suggesting local membrane destabilization despite the presence of dystrophin. Additionally, we confirmed the presence of a truncated form of both dystrophin isoforms Dp427, and for the first time Dp116, in DMD hiPSC-CMs and observed that dystrophin levels were comparable between DMD and repaired isogenic control cells (Fig. 4E). Of note, Dp427 dystrophin was absent in DMB03 DMD cardiomyocytes, with deletion of exons 48-49, while Dp116 was present, as mutation was before the promoter for Dp116 isoform (Fig. 4E). Furthermore, both immunofluorescence staining (Fig. 4D) and western blot analysis (Fig. 4F) confirmed the presence of dystrophin in hiPSC-SMs. Due to the cellular morphology, potential alterations in the subcellular localization of dystrophin and DAG1 in DMD hiPSC-SMs could not be clearly determined. However, western blot analysis revealed a reduced level of Dp427 isoform in DMD muscles compared to gene-repaired counterparts, as well as the absence of Dp116 isoform expression (Fig. 4F), consistent with its cell-specific expression reserved to Schwann cells, and as indicated by us, also to cardiomyocytes.

### 3.5 The effect of isoproterenol and metoprolol on electrophysiological properties of hiPSC-CMs

In the final step, we evaluated the β-adrenergic responsiveness of repaired and DMD hiPSC-CMs using microelectrode array (MEA) recordings. Cells were seeded on 24-well MEA plates, and spontaneous field potentials were recorded under basal conditions, followed by sequential application of isoproterenol (ISO, 1=µM) and metoprolol (MET, 1=µM). Under basal conditions, beating frequency (Fig. 5A), beat period (Fig. 5B), and field potential duration (FPD) (Fig. 5C) were comparable between groups. Upon ISO stimulation, beating frequency significantly increased in both groups, with DMD hiPSC-CMs exhibiting a markedly higher rate than repaired cells (p < 0.0001). In parallel, beat period and FPD were significantly prolonged in repaired hiPSC-CMs relative to DMD (p < 0.0001), indicative of altered repolarization dynamics. Following MET treatment, beating frequency decreased in both groups (p < 0.01), while FPD and beat period remained modestly prolonged in repaired hiPSC-CMs (p < 0.05). Spike amplitude remained comparable between groups across all conditions, with no statistically significant differences, although values tended to be marginally elevated in DMD cells (Fig. 5D). Representative MEA traces (Fig. 5E) recapitulate these findings, showing narrower spike morphology and shorter duration in DMD hiPSC-CMs, consistent with faster repolarization. In contrast, repaired cells displayed broader waveform profiles and longer repolarization phases, aligning with extended FPD measurements. Together, these data reveal subtle yet functionally relevant differences in electrophysiological behaviour between DMD and repaired cardiomyocytes under adrenergic stimulation.

**Fig. 5.**
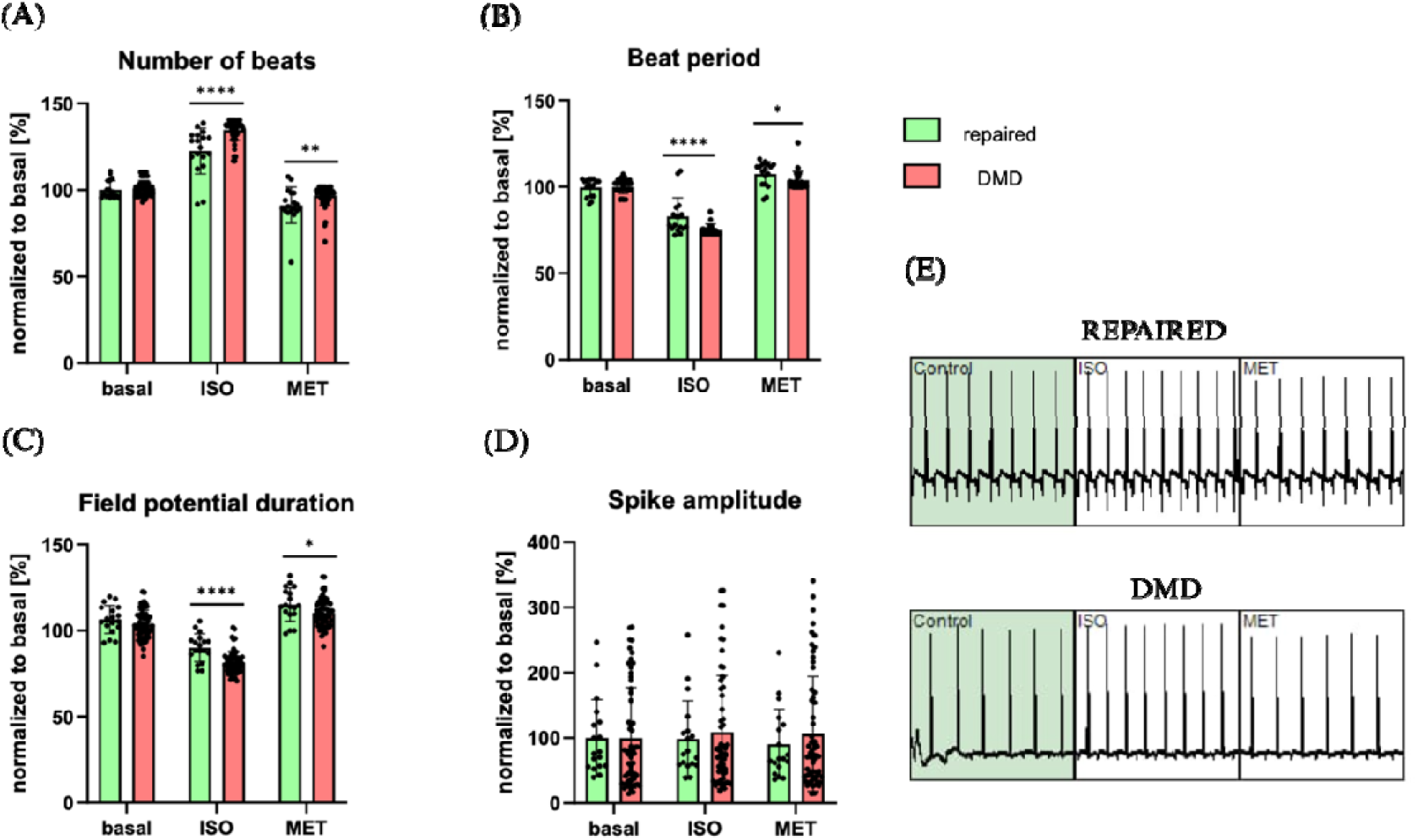
Electrophysiological profiling of repaired and DMD hiPSC-CMs. Analysis of **(A)** beat rate, **(B)** beat period, **(C)** field potential duration (FPD), and **(D)** spike amplitude under basal conditions, and following treatment with isoproterenol (ISO) or metoprolol (MET). The values obtained for basal, ISO, and MET recordings were then divided by the corresponding basal mean and expressed as a percentage (%). n=2. Unpaired Student’s *t*-test with normal distribution was used for the statistical analysis. **(E)** Overview of the last 10 seconds of multielectrode array (MEA) raw traces recorded from repaired and DMD hiPSC-CMs under basal conditions and following treatment with ISO or MET. hiPSC-CMs – human induced pluripotent stem cell-derived cardiomyocytes.

## 4. Discussion

Our study reports a previously uncharacterized in a DMD patient genetic variant c.9975-1G>T in intron 68 of *DMD* gene, which was expected result in the absence of both Dp427 and Dp116 dystrophin isoforms. In this research, we established a hiPSC line with described G>T transversion and used CRISPR/Cas9 technology for its precise correction by HDR (Fig. 1), creating a new tool for DMD modeling. Obtained isogenic repaired and DMD cell lines were characterized by normal karyotype, expressed markers specific for pluripotency and three germ layers (Fig. 2) and differentiated into both hiPSC-CMs and hiPSC-SMs (Fig. 3).

The mutation identified in the patient resides in the terminal nucleotide of intron 68, within the 3′ splice acceptor site, and was predicted to disrupt normal splicing, resulting in skipping of exon 69, introduction of a premature termination codon, and consequent loss of dystrophin expression. Surprisingly, transcript analysis revealed the presence of exon 69 in *DMD* mRNA in both repaired and DMD hiPSC-CMs and hiPSC-SMs (Fig. 4B). These results are inconsistent with the proposed mechanism of the effects of 3’ splice site mutations. According to the literature, such alterations should lead to either exon skipping or activation of a nearby cryptic splice site, resulting in pseudo exons formation and the inclusion of a sequence fragment into the transcript ^22–24^. Moreover, subsequent sequencing of dystrophin mRNA demonstrated, that c.9975-1G>T transversion caused an excision of only first six nucleotides of exon 69 (Fig. 4B). A similar case was reported in 2016 in a DMD patient carrying a 3’ splice acceptor site mutation in intron 26, which caused omission of only first two nucleotides of exon 27 rather than complete exon skipping. This frameshift generated a premature termination codon, resulting in a loss of dystrophin expression and DMD symptoms, including cardiomyopathy ^25^.

On the contrary, our findings indicate intriguing results: the deletion of two amino acids, tyrosine and arginine (positions 3325 and 3326) and preservation of the reading frame. Interestingly, dystrophin levels in repaired and DMD hiPSC-CMs were comparable (Fig. 4E), while a pronounced reduction in dystrophin expression was evident in DMD hiPSC-SMs (Fig. 4F), however truncated protein expression is characteristic not for DMD, but BMD patients ^26^. Nonetheless, there have been reports of patients carrying missense mutations with expression of dystrophin and yet severe DMD phenotype ^27^. Therefore, the symptoms leading to DMD diagnosis in the described patient could be resulting from dystrophin’s loss of function, as DMD hiPSC-CMs exhibited some abnormalities in the localization of both dystrophin and DAG1 (Fig. 4C).

The cysteine-rich domain encoded by exons 62–69 of the *DMD* gene consists of: WW domain, two EF hands and a ZZ domain, and plays a critical role in binding dystrophin to β-dystroglycan, whereas exons 69–79, which constitute the C-terminal domain, facilitate interactions with syntrophin and dystrobrevin ^28^. It has been reported that mutations causing changes in highly conserved ZZ domain, especially in the residues 3326-3332 of dystrophin, distort β-dystroglycan binding and lead to a severe muscle-wasting phenotype ^29^, even if only a single amino acid is deleted ^30^. A few years after initial diagnosis the patient described in our study has been treated with 18 mg deflazacort, exhibited elevated CK levels (11 778 U/l), hyperlordosis, calf hypertrophy, NSAA score of 28/34 and distance of 375 m in 6MWT. The patient remained clinically stable, without signs of respiratory failure or cardiomyopathy; nevertheless, the phenotype remained consistent with DMD, underscoring the functional importance of tyrosine 3325 and arginine 3326 within the dystrophin ZZ domain.

Dp116 is a dystrophin isoform characteristic of Schwann cells ^31^, however the presence of its transcript has also been detected in cardiac muscle ^7^. To best of our knowledge, we are the first to confirm Dp116 expression in hiPSC-CMs on the protein level (Fig. 4E). This key finding could provide valuable insights for further exploration of DMD-associated cardiomyopathy, as it has been reported that mutations in Dp116 were associated with milder cardiac involvement compared to mutations in other dystrophin isoforms ^7^.

MEA-based technologies offer a non-invasive approach to interrogate the electrophysiological properties of hiPSC-CMs, supporting drug screening and proarrhythmic risk assessment ^32^. To assess functional differences relevant to DMD cardiomyopathy, we compared DMD and repaired hiPSC-CMs under basal and adrenergic conditions. At baseline, no significant differences were observed in beating frequency, beat period, or field potential duration (FPD) (Fig. 5A–C), indicating preserved basic electrophysiological function in DMD cells. We next applied 1=µM isoproterenol (ISO), a non-selective β-adrenergic agonist that enhances cardiac contractility and heart rate by stimulating β-receptors ^33^. Both groups responded with increased beating frequency, though DMD hiPSC-CMs exhibited a stronger chronotropic response, with shorter beat period and FPD compared to repaired cells. These changes suggest altered repolarization dynamics, likely due to increased Ca²= influx through Piezo1 channels, modified SERCA2A activity ^33^, and sarcolemmal instability. Subsequent treatment with 1=µM metoprolol (MET), a selective β=-adrenergic antagonist that reduces heart rate and myocardial contractility ^34^, returned beating frequency toward baseline in both groups and prolonged FPD compared to ISO treatment. Spike amplitude was similar, with slightly elevated values in DMD cells (Fig. 5D). MEA traces (Fig. 5E) showed narrower, shorter spikes in DMD hiPSC-CMs, indicating faster repolarization, while repaired cells exhibited broader waveforms. Together, these findings reveal distinct yet functionally relevant electrophysiological differences in repaired vs. DMD hiPSC-CMs under adrenergic stimulation. It remains to be established whether this is due to protein dysfunction, its lower amount or proteasomal degradation as observed in case of *DMD* missense mutations ^35,36^.

In summary, we identified and characterized a previously unreported intronic DMD variant (c.9975-1G>T), allegedly associated with the loss of Dp427 and Dp116 isoforms. Unexpectedly, the mutation led to a subtle in-frame deletion of six nucleotides within the critical ZZ domain, rather than full exon skipping, suggesting preserved transcript integrity but impaired dystrophin function due to distortion of β-dystroglycan binding. By generating isogenic hiPSC-derived cardiomyocytes and skeletal muscle cells, we uncovered differential dystrophin expression and confirmed, for the first time, Dp116 protein presence in hiPSC-CMs. Functional electrophysiological profiling revealed altered repolarization dynamics and enhanced adrenergic responsiveness in DMD cardiomyocytes, providing new insights into mutation-specific mechanisms underlying DMD-associated cardiomyopathy. Altogether, our results point to a phenotypic overlap between Duchenne and Becker muscular dystrophies, indicating that certain splice-site mutations may complicate clinical classification and prognosis. These findings underscore the need for further investigation into the molecular mechanisms underlying atypical dystrophin expression and function.

## Data Availability

All data are included in the article and its supplementary materials.

## Acknowledgements

Part of the optical microscopy experiments: confocal imaging of the immunofluorescence-stained cells was performed at the Bioimaging Laboratory, which serves as an imaging core facility at the Faculty of Biochemistry, Biophysics, and Biotechnology JU. During the preparation of this work the authors used ChatGPT in order to check and correct language and readability. After using this tool/service, the authors reviewed and edited the content as needed and take full responsibility for the content of the publication. Some figures were created using BioRender.com.

## Sources of Funding

The work was supported by the Polish National Science Centre (NCN) grant MAESTRO [2018/30/A/NZ3/00412] to J.D. The multi-electrode array (MEA) system used for electrophysiological analyses was purchased with funds from the Polish National Science Centre (NCN) grant SHENG-2 [2021/40/Q/NZ3/00165] to J.D.

## Disclosures

The authors have no relevant disclosures.

## Notes

### Competing Interest Statement

The authors have declared no competing interest.

### Author Declarations

The study was approved by the Bioethical Committee of the Warsaw Medical University (approval no. KB/111/2019, approved on 10.06.2019; project title: Molecular mechanisms of heart failure in Duchenne and Becker muscular dystrophy).

